# Normative Benchmarks for the Parent-report Nationwide Quality of Life Scale (P-NQLS)

**DOI:** 10.64898/2026.04.16.26350886

**Authors:** Yinuo Liu, Eric A. Youngstrom, Elizabeth A. Nienaber, Mary A. Fristad

## Abstract

**Introduction:** The Nationwide Quality of Life Scale (NQLS) is a brief, mental-health focused quality of life (QoL) scale with seven items that are non-overlapping with symptom scales. We developed a parent version (P-NQLS), obtained national norms, and calculated psychometric properties for the P-NQLS.

**Methods:** Parents (*N*=2251) of children aged 6-18 years who were representative of the U.S. population on key demographics completed the P-NQLS along with measures of depression, suicidality, internalizing, externalizing, and attention symptoms. We assessed the P-NQLS’s factor structure through exploratory factor analysis (EFA) and evaluated its internal reliability and convergent validity. Age- and sex-specific norms were established using GAMLSS with BCPE distributions and P-spline smoothers, with percentile curves and tables (5^th^-95^th^) provided.

**Results:** EFA suggested a one-factor solution for P-NQLS in the national sample. The scale showed good internal consistency (Cronbach’s *α*=0.85). P-NQLS total scores (M±SD=20.7±4.7, range=0-28, higher scores indicate higher QoL) were negatively correlated (all *p*<.0001) with depression (Pearson’s *r*=-0.47), suicidality (*r*=-0.50), internalizing (*r*=-0.43), externalizing (*r*=-0.41), and attention (*r*=-0.37) symptoms. P-NQLS scores declined steadily with age in both sexes, with the most pronounced decreases (3-5 points) observed at lower percentiles (5^th^, 10^th^), suggesting greater age-related decline among children with lower baselines. Females scored slightly higher than males across most ages and percentile levels, though the differences were within one point.

**Conclusions:** The newly created P-NQLS, a 7-item parent-reported QoL scale with one underlying factor, demonstrates strong reliability and validity and has robust national norms for youth aged 6-18.

## Introduction

Quality of Life (QoL) is a multidimensional construct encompassing physical wellbeing, social relationships, perception of futures, and other life domains (Felce & Perry, 1995). QoL measurement is taking on increased importance in mental health research and clinical practice, particularly as the forthcoming Diagnostic and Statistical Manual, 6^th^ Edition (DSM-6) is projected to place greater emphasis on measuring contextual factors, including social drivers of health and QoL, when assessing individuals (Brauser, 2025). This emphasis is especially relevant to children given their critical developmental stage. However, existing measures of child QoL tend to prioritize health-related indicators while insufficiently capturing children’s social conditions and subjective well-being (Wallander & Koot, 2016).

Our group previously developed a QoL scale designed specifically for youth with mental health concerns. Key features of the Nationwide Quality of Life Scale (NQLS) include its brevity (seven items), distinction from symptom severity measures, focus on relevant mental health treatment goals rather than physical health status (Covarrubias & Fristad, 2025), and free access via public domain. An initial psychometric study demonstrated that the NQLS had two factors (school and global functioning), internal reliability, convergent validity, and sensitivity to change, supporting its utility among treatment-receiving youth.

Multi-informant assessment is considered best practice in child mental health evaluation, as parents provide important and distinct perspectives on their children’s functioning (De Los Reyes & Epkins, 2023). Parents observe children across home, school, and community contexts and are often regarded as more reliable reporters of children’s mental health symptoms than children themselves (Youngstrom et al., 2011). In the domain of health-related QoL measurement specifically, parent-child agreement ranges from poor to good depending on the scale and domain assessed (Upton et al., 2008), suggesting that parent and child reports are not interchangeable but reflect different perspectives that could enrich clinical assessment. To capture this nuanced parent perspective on child QoL, we developed a parallel parent form of the NQLS, the Parent-report NQLS (P-NQLS). The P-NQLS mirrors the content and structure of the youth self-report NQLS, with questions reworded to reflect a parent’s perspective on their child’s functioning.

In addition to developing the P-NQLS, we normed the scale using a nationally representative sample as part of a larger public domain measurement-based care norming project — BENCHMARK 2025 (osf.io/yxf8z). Normative benchmarks describe whether a given score reflects typical or atypical functioning relative to the general population and could inform clinical decision making as it relates to symptom severity or impairment (Achenbach, 2001). In the context of child mental health and general functioning, national norms are especially valuable for tracking individual progress against population-level, non-clinical reference points, informing treatment plans, and flagging meaningful deterioration or improvement.

In this study, we examined psychometric properties of the P-NQLS and generated national norms. We evaluated the factor structure, internal consistency, convergent validity, and generated age- and gender-specific norms using a continuous regression norming method to support clinical interpretation of P-NQLS scores.

## Methods

### Procedure

This study was approved by the Institutional Review Board at Nationwide Children’s Hospital. The survey agency YouGov administered selected measures to parents. YouGov has a global panel of over 29 million registered members with a significant portion in the U.S., enabling the company to conduct nationally representative surveys. YouGov collected and cleaned all responses and applied stratification based on key demographics (child sex, race, ethnicity, and parent education level) to align with the 2023 American Community Survey (ACS) one-year estimates, resulting in a sample representative of the U.S. population on key demographics.

### Participants

After providing informed consent, parents of 2,000 children aged 6-18 completed the P-NQLS along with other scales as part of a larger study (osf.io/yxf8z). Based on parent report, children were on average 11.4 years old (*SD*=3.4) and distributed across the 6-18 age range, with 47.2% of children being female, 74.9% White, 9% Black or African American, 3.7% Asian, and 16.8% Hispanic, Latino, or Spanish Origin. Nearly all parents (97.7%) had obtained a high school diploma, and 47.4% had obtained a bachelor’s degree or higher. Median household income was between $70,000 and $79,999 annually. Key demographics are summarized in Table 1 along with 2023 ACS estimates.

**Table 1.** Participant demographic characteristics ^a^Data from 2023 American Community Survey (ACS) 1-year estimates.

|  | Parent Sample<br>(N=2251) | 2023 ACS<sup>a</sup> |
| --- | --- | --- |
| **Child age** | 11.74 (3.40) |  |
| 6-7 years | 15.5% |  |
| 8-9 years | 14.7% |  |
| 10-11 years | 14.3% |  |
| 12-13 years | 19.3% |  |
| 14-15 years | 19.9% |  |
| 16-18 years | 16.4% |  |
| **Child sex** |  |  |
| Female | 47.5% | 50.5% |
| Male | 52.5% | 49.5% |
| **Race** |  |  |
| White | 75.3% | 60.5% |
| Black or African American | 8.8% | 12.1% |
| Asian | 3.7% | 6.0% |
| American Indian or Alaska Native | 1.7% | 1.0% |
| Native Hawaiian or Pacific Islander | 0.3% | 0.2% |
| Some other race | 4.7% | 7.4% |
| Multiple races selected | 4.0% | 12.8% |
| **Ethnicity** |  |  |
| Hispanic, Latino, or Spanish Origin | 16.4% | 19.4% |
| Not Hispanic, Latino, or Spanish Origin | 83.6% | 80.6% |
| **Parent/guardian education** |  |  |
| High school graduate or higher | 97.6% | 89.8% |
| Bachelor's degree or higher | 47.5% | 36.2% |
| **Median Household Income** | \$70,000-\$79,999 | \$77,719 |

<sup>a</sup>Data from 2023 American Community Survey (ACS) 1-year estimates.

### Measures

#### Parent-report Nationwide Quality of Life Scale (P-NQLS)

Adapted from the youth self-report NQLS (Covarrubias & Fristad, 2025), the P-NQLS is a 7-item scale asking parents to rate how satisfied they are with the child’s friends, family, grades, school in general, health, life, and anticipated future. Each item is scored on a five-point Likert scale (0 = very unsatisfied to 4 = very satisfied). Total scores range from 0 to 28, where higher scores indicate parent perception of better child QoL.

Parents also completed the following measures assessing child mental and behavioral health. The *Very Quick Inventory of Depressive Symptomatology* (VQIDS) is a 5-item measure of core depression symptoms, including sad mood, self-outlook, involvement, fatigue, and psychomotor slowing (Rush et al., 2003). The VQIDS demonstrates a single-factor structure, acceptable internal consistency, and similar level of sensitivity to symptom change (De La Garza et al., 2017). The *Concise Health Risk Tracking* (CHRT) is a 16-item measure of suicidal behaviors and risks, including propensity, impulsivity, irritability, and suicidal ideation (Trombello et al., 2023). A 14-item version has since been validated in adolescents with suicidality, demonstrating a strong correlation with depression severity and suicidal thoughts, as well as sensitivity to change (Mayes et al., 2018). The *Pediatric Symptom Checklist-17* (PSC-17) is an interdisciplinary screening tool used to flag emotional, behavioral, and attention concerns in youth (Gardner et al., 1999). The PSC-17 has demonstrated strong psychometric properties across diverse pediatric populations, including good sensitivity and specificity for identifying children in need of mental health services. As the VQIDS and CHRT do not have established parent-report versions, we adapted self-report versions for parent use.

##### Analysis

Exploratory Factor Analysis (EFA) using principal axis factoring with promax rotation was used to explore the P-NQLS’s factor structure. Factor retention criteria included eigenvalues, scree plot, and parallel analysis. Internal consistency was evaluated using Cronbach’s alpha; convergent validity was established via Pearson correlations between the P-NQLS and other measures.

Age- and sex-specific national norms for the P-NQLS were determined with Generalized Additive Models for Location, Scale, and Shape (GAMLSS; Rigby & Stasinopoulos, 2005). Guided by Timmerman et al. (2021), multiple candidate distributions were compared; the best-fitting model was selected based on the lowest Bayesian Information Criterion (BIC) values (see Supplemental Table 1). The final model used a Box-Cox Power Exponential (BCPE) distribution with P-spline smoothers to capture age-by-sex interactions in mean (*μ*) and age-dependent changes in variance (*σ*), while holding skewness (*ν*) and kurtosis (*τ*) constant. The worm plot (Buuren & Fredriks, 2001) confirmed adequate local fit. Based on the GAMLSS output, we estimated smoothed percentile curves and tables (5^th^, 10^th^, 25^th^, 50^th^, 75^th^, 90^th^, 95^th^) separately for males and females.

## Results

### Factor Structure

The Kaiser-Meyer-Olkin test indicated good sampling adequacy with overall MSA=0.84. The Bartlett’s test revealed significant correlations between scale items (*χ*^*2*^=6429.756, *p*<.001), supporting the data’s suitability for factor analysis. In the EFA, the first factor had an eigenvalue of 3.75, while all remaining factors had eigenvalues below 1. The scree plot showed a clear elbow after Factor 1, supporting retention of a single factor. Parallel analysis based on component eigenvalues also suggested a one-factor solution. Across all retention criteria examined, evidence consistently supported a unidimensional structure of the P-NQLS, indicating that the scale captures one underlying construct of parent perception of child QoL. All items loaded strongly onto the single factor, with loadings ranging from 0.61 to 0.79 (see Table 2).

**Table 2.** Factor loadings of P-NQLS from Exploratory Factor Analysis.

| P-NQLS Item | Factor 1 |
| --- | --- |
| Friends | 0.61 |
| Family | 0.63 |
| Grades | 0.64 |
| School in General | 0.73 |
| Health | 0.62 |
| Life | 0.79 |
| (Anticipated) Future | 0.72 |

**Note.** P-NQLS: Parent-report Nationwide Quality of Life.

### Reliability and Validity

The P-NQLS showed good internal consistency with Cronbach’s *α*=.85 and McDonald’s ω=.86, indicating that the scale consistently measures one underlying construct. The P-NQLS also demonstrated strong convergent validity through statistically significant and negative correlations with established measures on child psychopathology (see Table 3). Moderate associations were observed between P-NQLS and CHRT-16 total (*r*=-0.50), PSC-17 total (*r*=-0.49), and VQIDS total (*r*=-0.47), indicating that lower P-NQLS scores (i.e., less desirable QoL) reliably track with greater suicidal risks, emotional and behavior problems, and depressive symptoms. Among the CHRT subscales, Propensity (*r*=-0.47) and Irritability (*r*=-0.43) showed moderate correlations with the P-NQLS, while Impulsivity (*r*=-0.36) and Suicidal Thoughts and (*r*=-0.27) showed weak correlations. For the PSC subscales, Internalizing (*r*=-0.43) and Externalizing (*r*=-0.41) were moderately correlated with the P-NQLS, with Attention (*r*=-0.37) slightly lower. Overall, the uniformly significant (*p*<.001) and negative correlations (*r*=-0.27 to -0.50) between the P-NQLS and other measures provided strong evidence that the P-NQLS is a valid measure of child mental health and wellbeing.

**Table 3.**
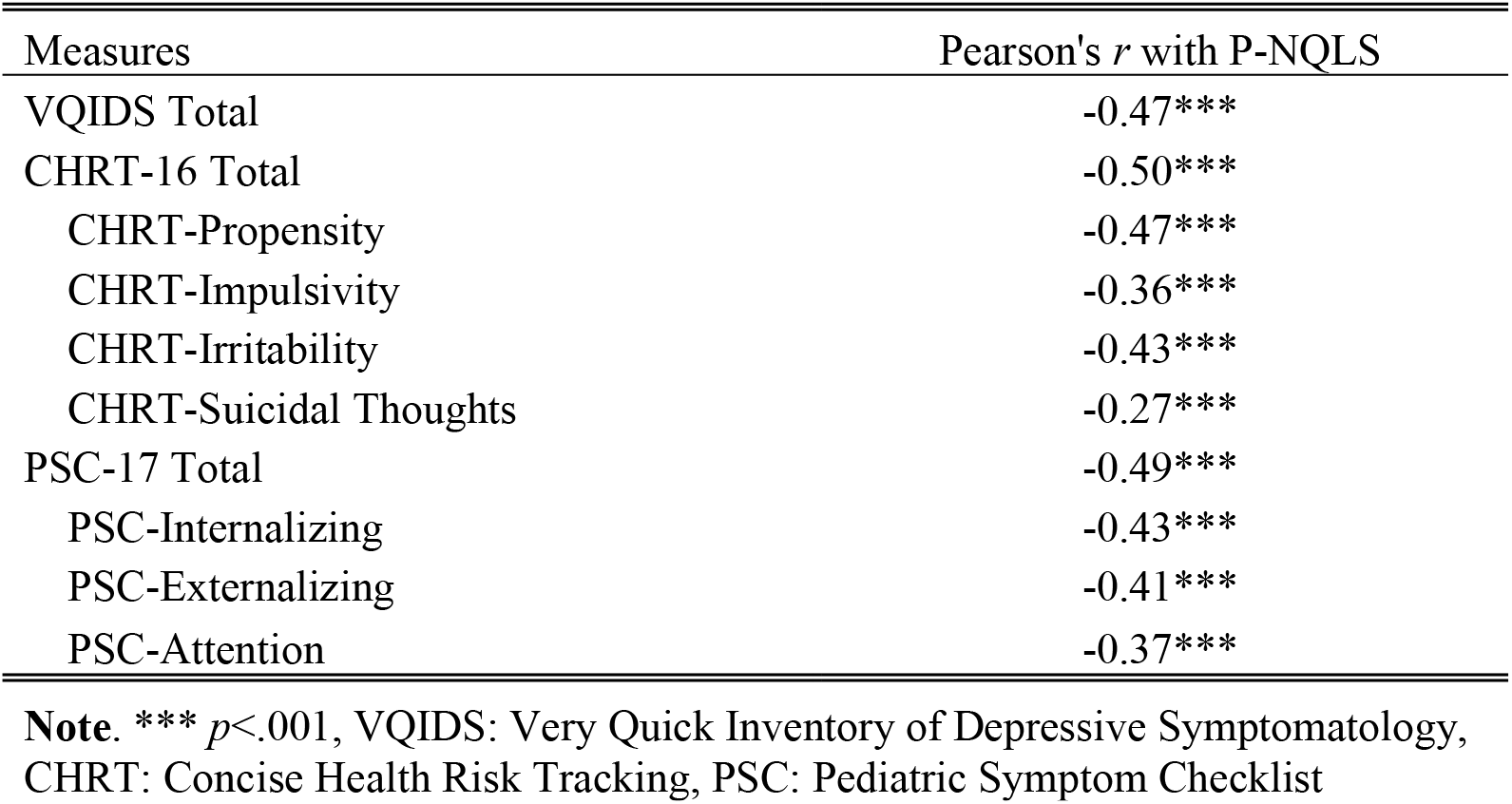
Pearson correlations between P-NQLS and other measures.

### National Norms

Percentile curves (see Figure 1) and tables (see Table 4) together revealed a consistent, albeit mild downward trend in P-NQLS total scores with increasing age across the 5^th^-95^th^ percentiles, regardless of child sex, suggesting that parental perception of child QoL steadily decreases as children grow older. For instance, the median (50^th^ percentile) P-NQLS score for males (Figure 1a) dropped from 22 at age 6 to 20 at age 18, and the median for females (Figure 1b) showed a drop of the same size from age 6 to 18.

**Table 4.** Key percentiles of Parent-report Nationwide Quality of Life (P-NQLS) total scores for males and females across 6-18 years.

| Male |  |  |  |  |  |  |  | Female |  |  |  |  |  |  |  |
| --- | --- | --- | --- | --- | --- | --- | --- | --- | --- | --- | --- | --- | --- | --- | --- |
| Age | P5 | P10 | P25 | P50 | P75 | P90 | P95 | Age | P5 | P10 | P25 | P50 | P75 | P90 | P95 |
| 6 | 14 | 16 | 19 | 22 | 25 | 26 | 27 | 6 | 15 | 16 | 19 | 22 | 25 | 27 | 28 |
| 7 | 14 | 16 | 19 | 22 | 24 | 26 | 27 | 7 | 14 | 16 | 19 | 22 | 25 | 27 | 28 |
| 8 | 13 | 16 | 19 | 22 | 24 | 26 | 27 | 8 | 14 | 16 | 19 | 22 | 25 | 27 | 28 |
| 9 | 13 | 15 | 18 | 22 | 24 | 26 | 27 | 9 | 13 | 15 | 19 | 22 | 25 | 27 | 28 |
| 10 | 13 | 15 | 18 | 21 | 24 | 26 | 27 | 10 | 13 | 15 | 18 | 22 | 25 | 27 | 28 |
| 11 | 12 | 15 | 18 | 21 | 24 | 26 | 27 | 11 | 13 | 15 | 18 | 22 | 24 | 26 | 27 |
| 12 | 12 | 14 | 18 | 21 | 24 | 26 | 27 | 12 | 12 | 15 | 18 | 21 | 24 | 26 | 27 |
| 13 | 12 | 14 | 17 | 21 | 24 | 26 | 27 | 13 | 12 | 14 | 18 | 21 | 24 | 26 | 27 |
| 14 | 11 | 14 | 17 | 21 | 24 | 26 | 27 | 14 | 12 | 14 | 18 | 21 | 24 | 26 | 27 |
| 15 | 11 | 13 | 17 | 21 | 23 | 25 | 27 | 15 | 11 | 14 | 17 | 21 | 24 | 26 | 27 |
| 16 | 11 | 13 | 17 | 20 | 23 | 25 | 26 | 16 | 11 | 13 | 17 | 21 | 24 | 26 | 27 |
| 17 | 10 | 13 | 17 | 20 | 23 | 25 | 26 | 17 | 11 | 13 | 17 | 21 | 24 | 26 | 27 |
| 18 | 10 | 13 | 16 | 20 | 23 | 25 | 26 | 18 | 10 | 13 | 17 | 20 | 24 | 26 | 27 |

**Fig. 1.**
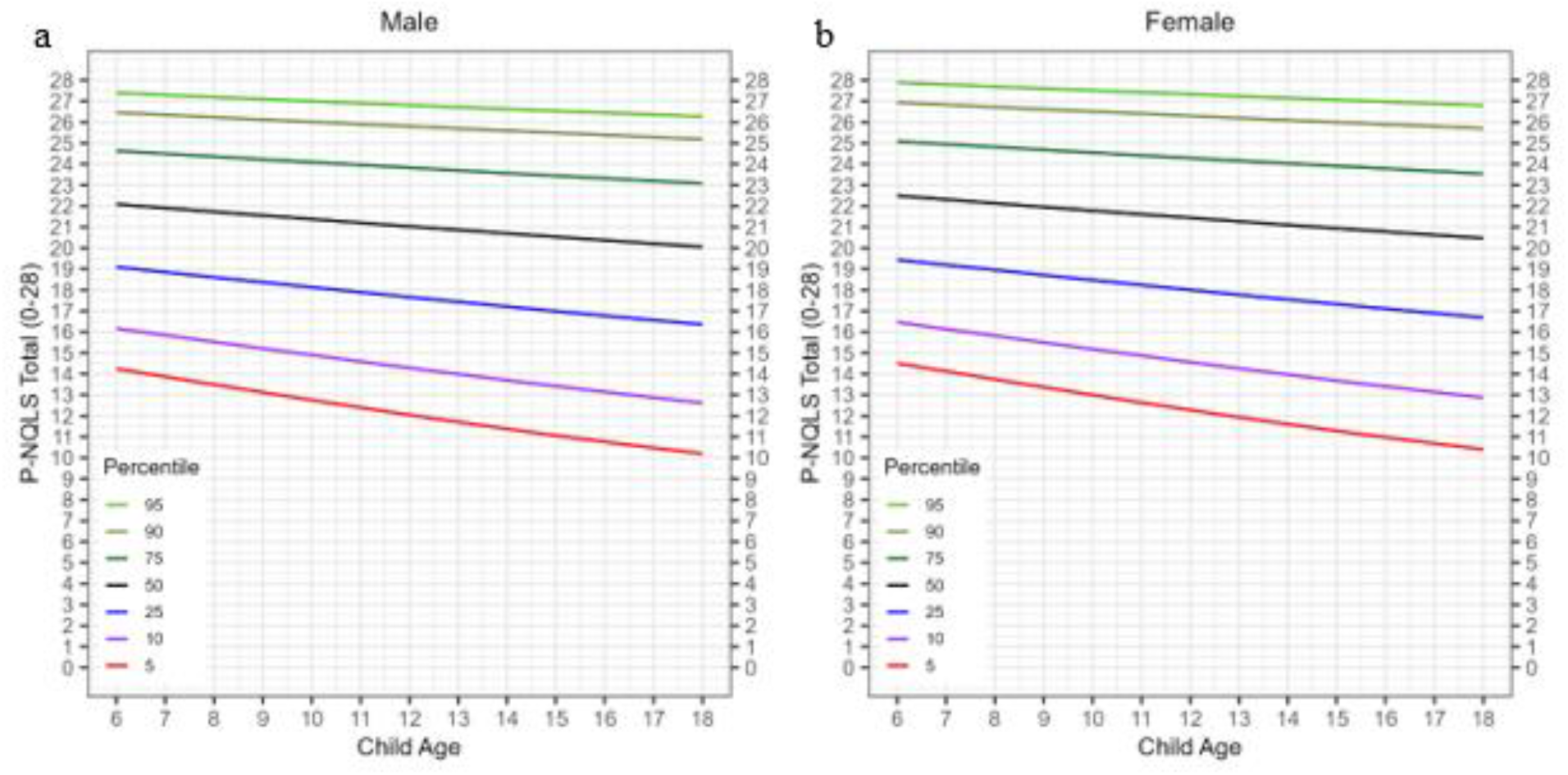
Percentile curves (5^th^-95^th^) for Parent-report Nationwide Quality of Life (P-NQLS) total scores (range: 0-28) across ages 6-18 (a. P-NQLS for males b. P-NQLS for females)

Notably, the magnitude of age-related decline was not uniform across percentile ranks, and lower percentiles exhibited steeper declines than upper percentiles. Among males, the 5^th^ percentile showed a 4-point change across the 6-18 age range (P-NQLS: 14→10), whereas the 95^th^ percentile showed a change of 1-point and remained at 26. A similar pattern was observed among females, with the 5^th^ percentile declining by 5 points (P-NQLS: 15→10) compared to the 95^th^ percentile declining by only 1 point (P-NQLS: 28→27). These findings indicated that children perceived by their parents as having lower QoL could be more severely affected by age-related stressors over time, widening the gap between higher- and lower-functioning children as they approach late adolescence.

A small but consistent sex difference was observed in P-NQLS total scores, with females scoring marginally higher than males across most age groups and percentile levels. Given that the difference remained within one point on a 0-28 scale, it may reflect measurement artifact rather than real-world disparities in QoL between sexes and should be interpreted with caution.

## Discussion

Measuring QoL, especially in children and adolescents, is increasingly important in measurement-based care. We previously demonstrated that the NQLS, a brief, youth self-report QoL measure, exhibited a two-factor structure (school and global functioning) with acceptable reliability, convergent validity, and sensitivity to change in a clinical sample (Covarrubias & Fristad, 2025). The present study extends those findings by introducing a parallel parent-report form, the P-NQLS, and establishing its psychometric properties and national norms in a large, nationally representative sample.

In this normative sample, the P-NQLS yielded a single-factor structure, contrasting the two-factor solution observed in the youth self-report NQLS. This difference suggests that parents perceive their child’s functioning as a more unified construct, without the degree of differentiation between school functioning and other life domains that youth self-report. Indices of internal consistency and convergent validity for the P-NQLS were largely comparable to those obtained for the NQLS in the clinical sample, supporting the psychometric robustness of this parent-report adaptation.

The normative data established in this study offer clinically meaningful benchmarks for interpreting parent-reported child QoL across development. By anchoring individual scores to a nationally representative reference sample, clinicians can move beyond raw scores to answer the question parents inevitably raise: “Is my child’s score normal?” Median P-NQLS total scores declined modestly but consistently with age, dropping two points from age 6 to 18 for both boys and girls. This pattern likely reflects developmental challenges associated with adolescence, including increasing academic demands (Lal, 2014), greater peer complexity (Buchanan & Bowen, 2008), and identity formation (Branje et al., 2021), all of which may erode parental perception of their child’s overall well-being. This phenomenon has also been captured by self-report measures, with an age-related decline in QoL reported in multiple cross-sectional studies (Goldbeck et al., 2007; Michel et al., 2009).

Importantly, the observed decline in reported QoL was not uniform across the P-NQLS score distribution. The gap between younger and older children was considerably more pronounced at the lowest percentiles (a 4-point decline for boys and a 5-point decline for girls at the 5^th^ percentile) compared to 1-point change at the 95^th^ percentile. This divergence suggests that children already perceived by their parents as having lower QoL may be disproportionately vulnerable to age-related stressors, potentially due to fewer psychological, social, or material resources to buffer against the challenges associated with adolescence. As a result, the gap between higher- and lower-functioning children widens with age, underscoring the importance of early identification and intervention for children showing early signs of QoL impairment. Additionally, QoL in adolescence and young adulthood can be predicted by early impairments in emotional and behavioral health domains such as emotion regulation, social skills, self-esteem, and self-control (Lal, 2014). By addressing both early QoL and behavior concerns, children can be set up for better longitudinal outcomes.

There are two major limitations with the current study. First, the P-NQLS has not been tested in a clinical sample and its sensitivity to change in response to treatment remains unknown. While clinicians can use the normative benchmarks to flag cases warranting closer attention or intervention, the percentile anchors do not establish diagnostic thresholds or capture treatment-related changes. Second, although YouGov recruited a nationally representative sample by stratifying on key demographics, some groups remained underrepresented relative to ACS benchmarks, including Black and multiracial children, Asian children, and families from lower educational and income backgrounds. This reflects an inherent limitation of panel-based survey recruitment, where certain populations are systematically harder to reach regardless of stratification efforts. The extent to which the current norms generalize to children from underrepresented racial, ethnic, and socioeconomic backgrounds warrants further investigation.

In conclusion, the P-NQLS is a brief, publicly available, and psychometrically sound parent-report measure of child QoL that is well-suited for use in mental health settings. The national norms established in this study provide clinicians with age- and sex-specific benchmarks to support score interpretation and measurement-based care across the full span of childhood and adolescence. Future studies should examine parent-child concordance and establish the scale’s utility in clinical samples.

## Data Availability

Code and sufficient data for replication available upon request from the corresponding author.

## Authors’ Contributions

EAY conceptualized the project and provided data; YL ran all analyses and wrote methods and results; MAF wrote introduction and discussion; EAN formatted the manuscript; all authors reviewed and edited the full manuscript.

## Ethics Committee Approval

The Institutional Review Board of Nationwide Children’s Hospital provided a Non-Human Subject Research determination for analyses of deidentified survey data.

## Data Access Statement

Code and sufficient data for replication available upon request from the corresponding author.

## Acknowledgments/Role of Funding Source

Supported by a gift from the Chlapaty family to Nationwide Children’s Hospital to support the Institute for Mental and Behavioral Health Research (IMBHR).

---

**Supplemental Table 1.** GAMLSS Model fit indices of candidate distributions.

| Model Distribution | BIC |
| --- | --- |
| **BCPE** | **13139.99** |
| BCCG | 13144.28 |
| Normal | 13346.81 |
| T-Family | 13306.29 |

